# Evaluation of sampling and concentration methods for *Salmonella enterica* serovar Typhi detection from wastewater

**DOI:** 10.1101/2022.06.07.22275929

**Authors:** Nicolette A. Zhou, Angelo Q.W. Ong, Christine S. Fagnant-Sperati, Joanna Ciol Harrison, Alexandra L. Kossik, Nicola K. Beck, Jeffry H. Shirai, Elisabeth Burnor, Rachael Swanstrom, Bethel Demeke, Suhani Patel, Typhoid Environmental Surveillance Working Group, John Scott Meschke

## Abstract

*Salmonella enterica* serovar (*Salmonella* Typhi) is the causative bacterial agent of Typhoid fever. Environmental surveillance of wastewater and wastewater-impacted surface waters has proven effective in monitoring various pathogens, and has recently been applied to *Salmonella* Typhi. This study evaluated eight sample collection and concentration methods with twelve variations currently being developed and used for *Salmonella* Typhi surveillance globally to better understand the performance of each method based on their ability to detect *Salmonella* Typhi and feasibility. *Salmonella* Typhi strains, Ty21a and Ty2, were seeded to influent wastewater at known concentrations to evaluate the following methods: grab sampling using electropositive filters, centrifugation, direct enrichment, or membrane filtration and trap sampling using Moore swabs. Concentrated samples underwent nucleic acid extraction and were detected and/or quantified via qPCR. Results suggest that all methods tested can be successful at concentrating *Salmonella* Typhi for subsequent detection by qPCR, although each method has its own strengths and weaknesses including the *Salmonella* Typhi concentrations they are best suited for with a range of positive detections observed as low as 0.1-0.001 CFU Ty21a/mL and 0.01 CFU Ty2/mL. These factors should be considered when identifying a method for environmental surveillance and will greatly depend on the use case planned.

## INTRODUCTION

The World Health Organization (WHO) estimates the annual global death toll from typhoid fever to be between 128,000-161,000 people [1]. The pathogen responsible for typhoid fever is the gram-negative bacterium *Salmonella enterica* serovar Typhi (*Salmonella* Typhi). Humans are the only known natural host and reservoir for *S*. Typhi, which is spread fecal-orally through water, food, or objects contaminated with feces of an infected individual [2]. Typhoid fever is a human health risk particularly in low and middle income countries where access to clean water and adequate sanitation can be a challenge [3,4]. Typhoid fever is endemic in most South Asian and Sub-Saharan African countries, however incidence varies by season and localized outbreaks can occur [5,6]. While the annual estimated number of cases ranges between 11-21 million people [1], this global burden estimate is based primarily on limited clinical blood culture surveillance data. However, surveillance based on diagnostic clinical microbiology is expensive, requires specialized facilities, is approximately 50-60% sensitive, and depends on symptomatic individuals presenting to healthcare facilities and on clinicians performing a diagnostic test [7]. Often clinical treatment is not sought [8,9]. Additionally, asymptomatic carriers or pauci-symptomatic individuals do not tend to have diagnostic tests performed though these cases can still shed the pathogen in feces [6]. For these reasons, global typhoid burden is likely underestimated [10].

Given the challenges with conventional approaches to typhoid surveillance, novel strategies are necessary. One strategy that has proven effective in monitoring pathogens is environmental surveillance (ES). ES is the collection of soil, water, air or other environmental samples, and analysis for pathogens. Since *Salmonella* Typhi is shed through feces, the organism is expected to be in wastewater or wastewater-impacted surface waters of locations with outbreaks or endemic transmission [11,12]. ES for *Salmonella* Typhi may inform on disease burden in the population and help identify typhoid hot spots. ES has been shown to support the reduction of diseases and prevention of outbreaks of various enteric pathogens [13–16]. The WHO first recommended Typhoid Conjugate Vaccine (TCV) in March 2018 [17], but introduction has been limited to locations at highest risk and greatest burden due to limited availability and funding options. Thus, ES can be used to help guide TCV deployment decisions, and later to monitor effectiveness of intervention strategies such as TCV introduction.

Multiple collection and concentration methods have recently been developed and are applied globally for downstream analysis to detect *Salmonella* Typhi in water sources, and this study will focus on wastewater [12]. ES methods for pathogen surveillance use grab sampling or trap sampling. These techniques generally avoid collecting large sediments by sampling the wastewater or wastewater-impacted water surface, or pre-filtering the water through a coarse filter. The volumes typically processed in grab sample methods vary from 50 milliliters to 20 liters [18–22], and samples are collected from outlet pipes, open sewer channels, surface waters, or other sewage stream. Grab samples are either concentrated in-field via gravity filtration, or are transported to a laboratory with proper cold chain for subsequent concentration, elution, and/or enrichment [10,18,23–28]. Sample concentration enables a larger volume of wastewater or wastewater-impacted water to be assayed, which is important when low concentrations are anticipated such as is typically the case in environmental samples. Methods for sample concentration also vary – some filter samples through an electropositive ViroCap filter, 0.45-μm membrane filters, or ultrafilters; other methods include centrifugation [12,18–22,26,29]. Trap sample methods for *Salmonella* Typhi have historically utilized a Moore swab method [30,31]. The Moore swab, or Moore cotton tampon is suspended into the water source for up to 6 days [29,32–37] to allow for bacteria flowing through water to adsorb on the swab, but not for large debris to get trapped. Swabs are then enriched using different growth/enrichment mediums. After processing, samples are analyzed using culture-based detection methods and/or quantitative polymerase chain reaction (qPCR). Although culture-based methods have historically been used to isolate *Salmonella* Typhi, recovery has been inconsistent and culturing is challenging; therefore, qPCR is increasingly used for detection of *Salmonella* Typhi in wastewater [10,29,38]. Further, when enrichment is not used in sample processing (unless most probable number methodologies are employed), qPCR enables quantification of the organism’s concentration in the original sample and allows for detection of viable but not culturable *Salmonella* Typhi, whereas culture confirms viability of organism and permits comparative genomic analysis.

Of the methods used for sampling and detection of *Salmonella* Typhi from environmental wastewater and wastewater-impacted surface water samples, none would be uniformly appropriate as a stand-alone method for all study designs, sampling locations, and wastewater matrices. The aim of this study was to evaluate various sample collection and concentration methods currently being developed and used for *Salmonella* Typhi surveillance globally to better understand the performance of each method based on their ability to detect *Salmonella* Typhi and feasibility. Eight methods with twelve variations were evaluated in total for detection of *Salmonella* Typhi in a wastewater matrix. Additional ES methods for *Salmonella* Typhi including dead-end ultrafiltration and hollow fiber ultrafiltration were not able to be included in this study due to time and resource constraints. The methods evaluated consisted of grab sampling using electropositive filters, centrifugation, direct enrichment, or membrane filtration and trap sampling using Moore swabs. Concentrated samples underwent nucleic acid extraction and were detected and/or quantified via qPCR. The detection of *Salmonella* Typhi, each method’s feasibility (active time, total time, safety, key processing supplies per sample, and key lab equipment), and potential use cases were also evaluated.

## METHODS

### Organism culture and enumeration

Two commonly used strains of *Salmonella* Typhi were utilized as positive controls in this study, Ty21a and Ty2. *Salmonella* Typhi strain Ty21a is used in the oral, live attenuated typhoid vaccine and is negative for the Vi antigen. *Salmonella* Typhi strain Ty2 is a well-characterized, reference strain that is positive for the Vi antigen. To confirm presence of the Vi antigen in Ty2 prior to experiments, a Vi antigen agglutination test was performed using the Difco™ Salmonella Vi Antiserum (Becton, Dickinson and Company, Franklin Lakes, NJ, USA). *Salmonella enterica* serovar Typhimurium (*Salmonella* Typhimurium) was used as a negative control in the agglutination test. Ty21a (ATCC® 33459), Ty2 and *Salmonella* Typhimurium were obtained from Dr. Stephen Libby (University of Washington). For each strain, 10 µL antiserum and 10 µL overnight culture were combined on a glass slide and examined for agglutination after 15 minutes. Agglutination indicated the presence of the Vi antigen in Ty2.

Throughout this work, Ty21a was grown using LB-Miller broth (IBI Scientific, Dubuque, IA, USA) and Ty2 was grown in the dark using LB-Miller broth with a supplemental aromatic amino acid mix and 50 ng/mL ferrioxamine E (Millipore, Burlington, MA, USA). The aromatic amino acid mix was prepared in a 100×stock consisting of L-Phenylalanine (4 mg/mL) (TCI America™, Portland, OR, USA), L-Tryptophan (4 mg/mL) (Acros Organics, Fair Lawn, NJ, USA), 2,3-Dihydroxybenzoic Acid (1 mg/mL) (TCI America™), and Para-Aminobenzoic Acid (1 mg/mL) (Sigma Aldrich, St. Louis, MO, USA), which were dissolved in deionized water and filter sterilized. To improve our understanding of Ty21a and Ty2 growth and therefore ensure experiments were seeded during exponential growth, growth curves were determined for Ty21a and Ty2 and measured via optical density at 600 nm (OD_600_) and spot plating 100 µL of relevant dilutions [39] on LB-Miller agar, or LB-Miller agar with a supplemental aromatic amino acid mix and 50 ng/mL ferrioxamine E. Ty21a and Ty2 inocula for experiments were prepared by growing the organisms for a specified period of time at 37°C, monitoring for exponential growth phase, and storing single-use aliquots of the cultures in 30% glycerol until use (-80°C). Prior to planned experiments, 20 µL of the frozen Ty21a or Ty2 glycerol stock was inoculated in 15 mL of liquid media and incubated with shaking (200 rpm, 37°C, 12-16 hours).

### Study design

Primary influent wastewater grab samples were collected from a local wastewater treatment plant in Seattle, WA, USA, which processes 90 million gallons per day (mgd) during the dry season and can process over 300 mgd during the rainy season [40], resulting in matrix variability. Grab samples (7 to 14 L per carboy) were stored at 4°C until processing (conducted within 72 hours of collection). Varying amounts of Ty21a or Ty2 were seeded to 10 mL 1×phosphate-buffered saline (PBS), vortexed (30 seconds), and seeded into varying volumes of wastewater to reach the target concentrations. The final concentration of Ty2 or Ty21a in the seeded wastewater varied depending on experiment and ranged from approximately 0.001 to 10,000 colony forming units (CFU) per milliliter (Figure S1). Methods tested at the same concentration level primarily utilized the same initial wastewater matrix with replicates of three or six to enable comparison between the methods. The seeded concentrations were assessed in parallel for each experiment via spread plating of 100 µL of relevant dilutions on LB-Miller agar (Ty21a) or LB-Miller agar with a supplemental aromatic amino acid mix and 50 ng/mL ferrioxamine E (Ty2). The seeded wastewater was thoroughly mixed and then distributed using a peristaltic pump while continuously shaking for processing by *(1)* filter cartridge, *(2)* differential centrifugation, *(3)* grab enrichment, *(4)* membrane filtration, and *(5)* Moore swab methods (Figure 1). The methods were evaluated in a laboratory setting, and factors considered included: feasibility of performing these methods in other settings and *Salmonella* Typhi detection by qPCR. The feasibility was based on processing time, equipment, and supplies needed. Active time was defined as the total hands-on time required to conduct an individual method, excluding time in which personnel can complete other tasks. These periods may include incubation, shaking, and filtration.

**Figure 1.**
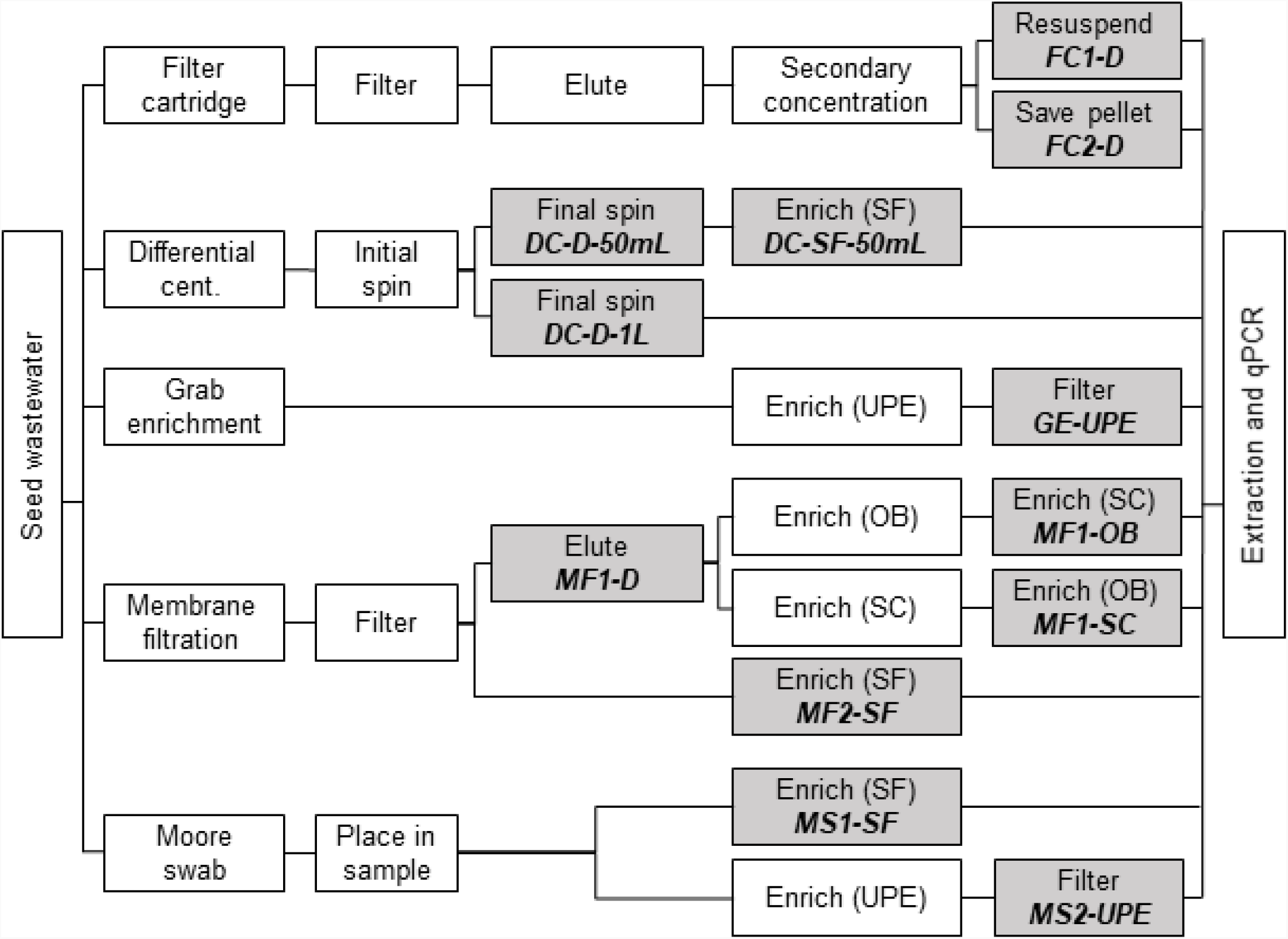
Workflow of method evaluations. Shaded boxes indicate where samples were collected for DNA extraction. D is direct; SF is selenite F broth; UPE is universal pre-enrichment broth; OB is ox bile; SC is selenite cystine broth; and qPCR is quantitative Polymerase Chain Reaction. Italicized, bolded text indicates method names.

### Methods evaluated

#### Two-inch filter cartridge method

Two variations of the two-inch filter cartridge method were tested, mentioned hereafter as FC1-D and FC2-D. The main difference between the two methods was the input for DNA extraction (half of the resuspended pellet vs. the full sample pellet) and the resulting difference in effective volume assayed (Table 1, Supplementary Information) [20,21,41].

**Table 1.** Volumes processed and effective volume entering qPCR for each method evaluated

| Method type | Method name | Enrichment | Initial volume | Average volume processed ($\pm 95\%$ CI) | Average effective volume entering qPCR<sup>a</sup> ($\pm 95\%$ CI) | Organism tested |
| --- | --- | --- | --- | --- | --- | --- |
| Filter cartridge | FC1-D | N/A | 6 L | 5.6±0.6 L | 85±22 mL | Ty21a |
| | FC2-D | N/A | 6 L | 4.6±0.5 L | 123±27 mL | Ty21a, Ty2 |
| Differential centrifugation | DC-D-50mL | N/A | 50 mL | 50 mL | 1.7±0.3 mL | Ty21a |
| | DC-SF-50mL | SF | 50 mL | 50 mL | 0.145±0.022 mL | Ty21a |
| | DC-D-1L | N/A | 1 L | 1 L | 9.7±2.1 mL | Ty2 |
| Grab enrichment | GE-UPE | UPE | 20 mL | 20 mL | 0.104±0.016 mL | Ty21a |
| Membrane filtration | MF1-D | N/A | 1 L | 423±19 mL | 2.0±0.2 mL | Ty21a, Ty2 |
| | MF1-OB | OB to SC | 1 L | 440±28 mL | 0.112±0.014 mL | Ty21a |
| | MF1-SC | SC to OB | 1 L | 465±28 mL | 0.124±0.015 mL | Ty21a |
| | MF2-SF | SF | 1 L | 418±33 mL | 2.3±0.5 mL | Ty21a |
| Moore swab | MS1-SF | SF | 5 L | N/A<sup>b</sup> | N/A<sup>b</sup> | Ty21a, Ty2 |
|  | MS2-UPE | UPE | 5 L | N/A<sup>b</sup> | N/A<sup>b</sup> | Ty21a, Ty2 |

<sup>a</sup> Calculated values assume no benefit from enrichment and no losses throughout the concentration process
<sup>b</sup> Moore swabs volume not quantifiable as the processed volume is unknown
95% CI is the 95% confidence interval; qPCR is quantitative polymerase chain reaction; SF is selenite F; UPE is universal pre-enrichment; OB is ox bile; and SC is selenite cystine.

#### Differential centrifugation

Three versions of differential centrifugation methods were used: DC-D-50mL, DC-SF-50mL, and DC-D-1L. All methods involved centrifugation for 1 minute at 1000 ×*g*, 4°C, followed by transferring the supernatant centrifuging again (15 minutes, 4000 ×*g*, 4°C). Additional details are provided in the Supplementary Information.

#### Grab Enrichment

For grab enrichment (hereafter, GE-UPE), 20 mL of seeded wastewater were added to 180 mL of Universal Pre-Enrichment broth (UPE; Becton, Dickinson and Company) and incubated at 37°C (24 hours). After incubation, 20 mL of the enriched sample was membrane filtered through one mixed cellulose ester (MCE) filter (0.45 µm pore size, 47 mm diameter; Millipore). The membrane filter was cut into 6-10 pieces using sterile scissors, placed in a 2 mL screw top tube and stored at -20°C prior to DNA extraction.

#### Membrane filtration methods

Multiple variations of vacuum membrane filtration were tested: MF1-D, MF1-OB, MF1-SC, and MF2-SF (Fig. 1). MF1 methods were pre-filtered using a paper, cone-shaped coffee filter placed on top of a filtration cup with a MCE filter, while the MF2 methods did not involve pre-filtration. Additional details are provided in the Supplementary Information.

#### Moore swab methods

Two Moore swab methods were tested, with one enriched using Selenite F (SF) broth (hereafter, MS1-SF) and another using UPE broth (hereafter, MS2-UPE) (Supplementary Information).

### DNA extraction and qPCR

DNA extraction was performed on the samples using the QIAamp PowerFecal Pro DNA Kit (Qiagen, Hilden, Germany) according to manufacturer’s instructions, with the following modifications. The input was pelleted 1 mL aliquots of the samples (FC1-D, DC-D-50mL, DC-SF-50mL, DC-D-1L, MF1-D, MF1-OB, MF1-SC, MF2-SF, and MS1-SF), pelleted secondary concentrates (FC2-D), or sliced membrane filters (GE-UPE and MS2-UPE). The DNA was eluted in 60 µL and aliquoted into two 30 µL volumes or eluted in 120 µL and aliquoted into three 40 µL volumes, and stored at -20°C prior to qPCR.

Samples were analyzed for Ty21a and Ty2 via a qPCR assay targeting the *staG* gene commonly used for *Salmonella* Typhi detection in human clinical samples [38]. The reaction was carried out with 0.4 µM of the forward (5’-CGCGAAGTCAGAGTCGACATAG-3’) and reverse (5’-AAGACCTCAACGCCGATCAC-3’) primers, 0.15 µM of the probe (5’-FAM-CATTTGTTCTGGAGCAGGCTGACGG-BHQ), and 1× iTaq Universal Probes Supermix (Bio-Rad Laboratories, Inc., Hercules, CA, USA). A DNA extract input volume of 5 µL was used with a 25 µL reaction. Each DNA extract was analyzed undiluted and at a 10-fold dilution to monitor for PCR inhibition effects. Unseeded wastewater controls, no template controls, and a positive control dilution series (standard curve) of Ty21a or Ty2 were analyzed with each qPCR run. Unseeded wastewater controls were positive in 17 of 56 wells throughout the study with a minimum Ct value of 34.1 and maximum Ct value (considered positive) of 39.0. No template controls were not detected throughout the course of the study. Standard curve efficiencies averaged 104% and R-squared values were greater than 0.96. The input for the standard curves were prepared by centrifuging (10 minutes, 10,000 ×g) a 1-mL volume of the Ty21a or Ty2 overnight culture harvested during exponential growth (quantified via spread plating of 100 µL of relevant dilutions on LB-Miller agar [Ty21a] or LB-Miller agar with a supplemental aromatic amino acid mix and 50 ng/mL ferrioxamine E [Ty2]), removing the supernatant, and performing DNA extraction as described above. All samples and controls were tested in duplicate or triplicate.

Samples positive for *Salmonella* Typhi were defined as those that amplified with a Ct of 40 or lower in one or two of the two technical replicates with an appropriate shaped curve. Samples with a Ct greater than 40 were assumed to be negative due to the potential for spurious artifacts to interfere with detection. The limit of detection (LOD) was determined to be a Ct of 37 (corresponding to approximately 100 CFU/mL of the original culture) for both Ty21a and Ty2, and was defined as 95% samples positive for this assay on this qPCR instrument utilized (Bio-Rad CFX96 Instrument; Bio-Rad Laboratories, Inc.) [42]. The limit of quantification (LOQ) was determined to be a Ct of 33 (corresponding to approximately 1,000 CFU/mL of the original culture) based on a coefficient of variation below 35% [43]. As samples above the LOD but below a Ct of 40 have a higher likelihood of false negatives, samples were determined to be positive if they amplified at Ct of 40 or lower.

## Data analysis

Methods were primarily evaluated for their rate of *Salmonella* Typhi positivity. This was determined irrespective of the application of enrichment steps and the sample volume processed as larger sample volumes do not inherently yield greater positivity. The effective volume assayed for the various methods was determined though, as shown below. Methods were not able to be evaluated for recovery efficiency due to the use of enrichment steps in several methods and the use of Moore swabs (collecting an unknown amount of volume).

The concentration factor (Eq 1) and effective volume assayed were calculated (Eq 2) using Microsoft Excel 2016.

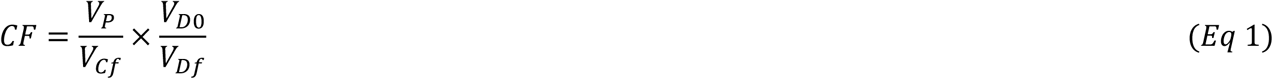

where *CF* is the concentration factor, *V*_*P*_ is sample volume processed and *V*_*Cf*_ is the final concentrate volume prior to DNA extraction, *V*_*D*0_ is the volume entering DNA extraction, and *V*_*Df*_ is the final concentrate volume after DNA extraction.

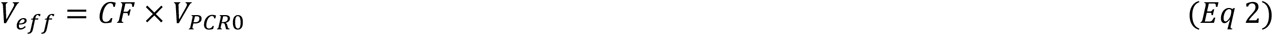

where *V*_*eff*_ is the effective volume assayed, and *V*_*PCR*0_ is the volume entering PCR reaction.

## RESULTS

### Method feasibility

The feasibility of these methods depends on a variety of factors such as timing, volumes concentrated, supplies and equipment required, and safety (Tables 1 and 2). The active time for each method is less than three hours. DC-D-50mL, DC-SF-50mL, GE-UPE, and MS1-SF methods each requires approximately 60 minutes or less of active personnel time. The total time to yield concentrates for DNA extraction can be obtained for most methods within 48 hours, including active and inactive time. The exception to this timing was for Moore swabs, which require 3 to 6 days for the concentrate due to the swab holding time in wastewater and the incubation time. The sample volume concentrated affects the processing time, with larger volumes requiring more processing time *(i*.*e*., filter cartridge, DC-D-1L, and membrane filtration). Procurement of supplies is simple as all required items are accessible commercially in the United States, though this may vary in other locations. For example, selenite-based enrichment broths can be difficult to obtain in other countries due to chemical safety concerns. These selenite-based enrichment broths are used in DC-SF-50mL, MF1-OB, MF1-SC, MF2-SF, and MS1-SF methods. Supply and equipment costs vary between methods and depend heavily on already existing supplies and equipment in other lab settings.

**Table 2.**
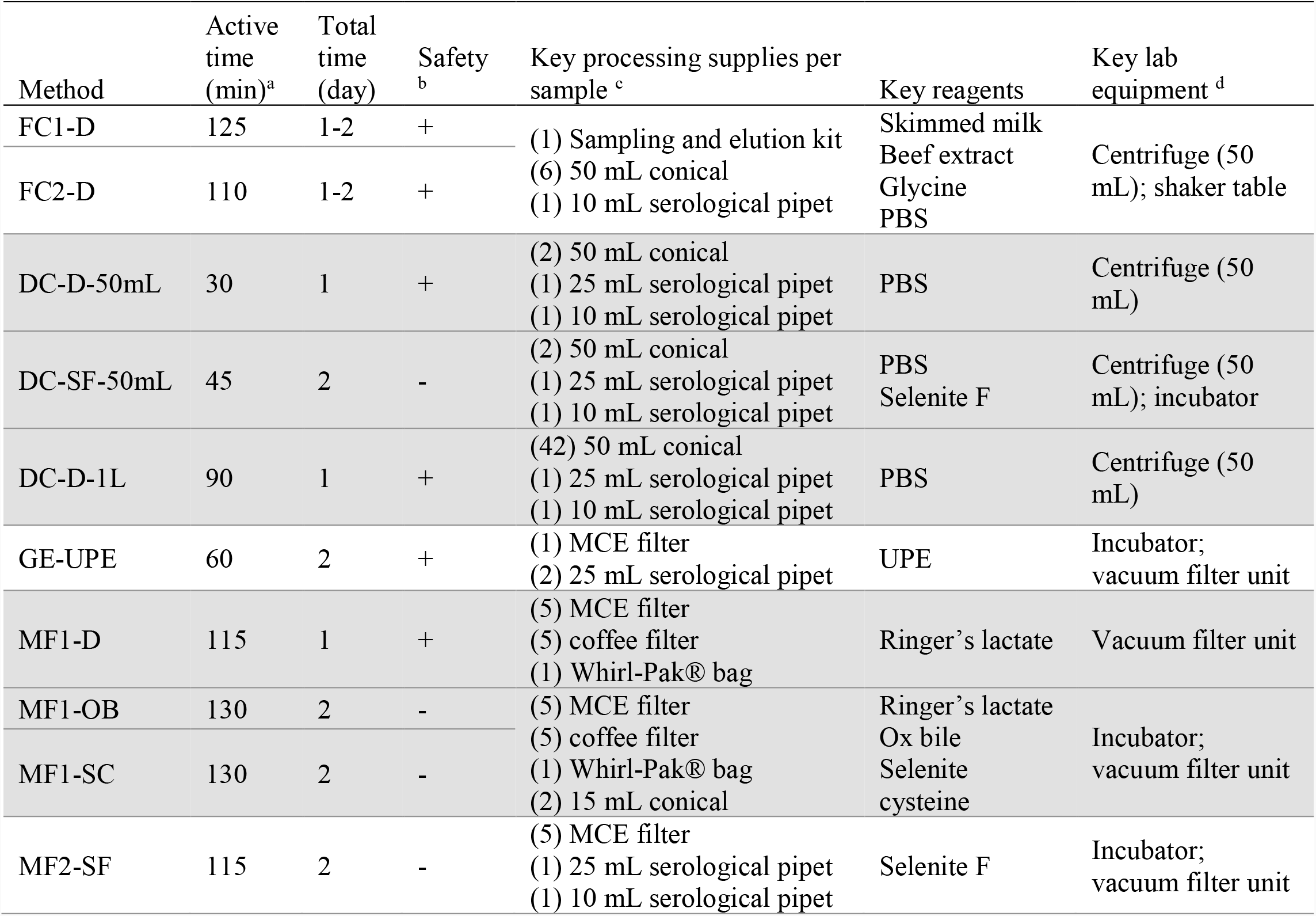
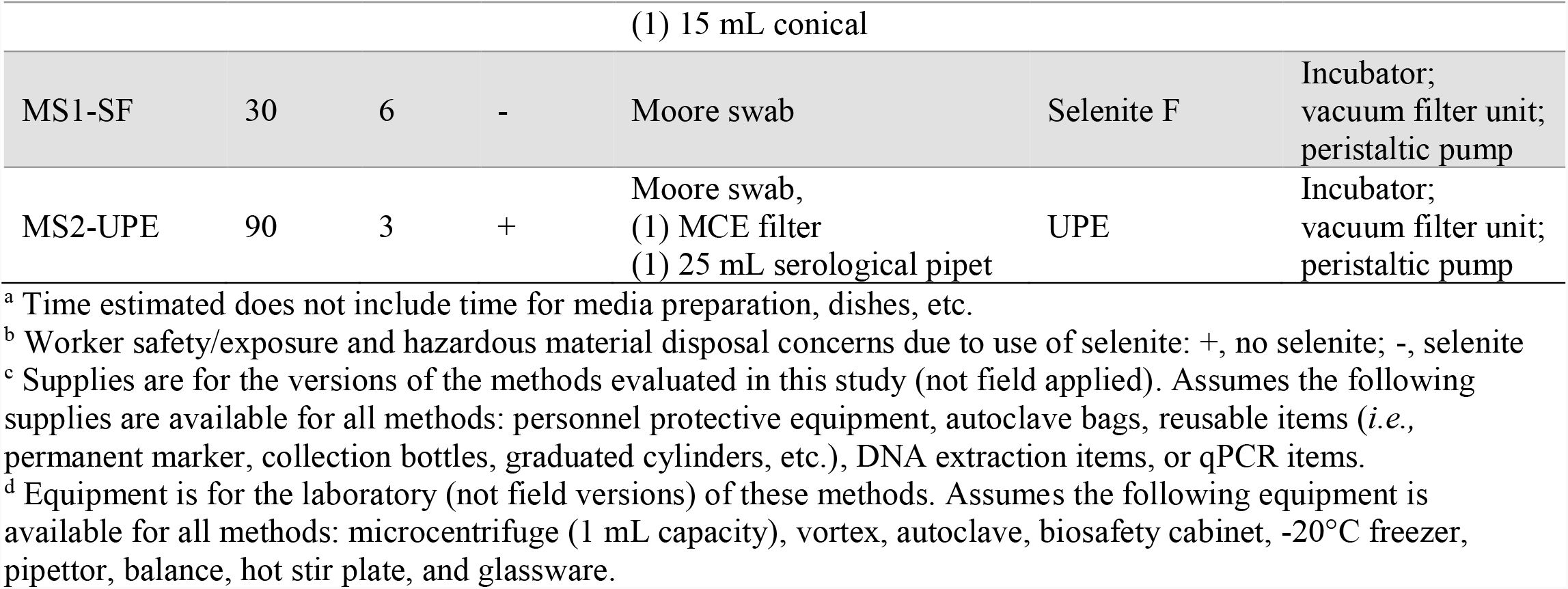
Details of collection and concentration methods tested effecting feasibility

### Ty21a detection

Eight methods with eleven variations total were tested for their ability to concentrate wastewater seeded with varying concentrations of Ty21a for detection via qPCR (Table 1).

When seeded at high concentrations (10,000 and 100 CFU/mL), all methods consistently detected Ty21a (100%; Table 3). Four methods maintained a high frequency of positive detection (>75%) at all concentrations tested: FC1-D, FC2-D, MF1-D, and MS1-SF, while other methods yielded fewer positive detections at lower seeded Ty21a concentrations. For DC-D-50mL, a high frequency of positive detection (100%) was maintained to a seeded concentration of 0.1 CFU/mL, compared to DC-SF-50mL samples which were minimally detected at or below a seeded concentration of 1 CFU/mL. Positive detection in GE-UPE samples also decreased at a seeded concentration of 1 CFU/mL. MF1-OB samples maintained a high frequency of positive detection to 0.01 CFU/mL, compared to MF1-SC samples which maintained a high frequency of positive detection to 0.1 CFU/mL. MF2-SF and MS2-UPE samples yielded similar results with a high frequency of positive detection down to 0.01 CFU/mL. Of the seven methods that maintained a high rate of positivity (>75%) at a seeded concentration of 0.01 CFU/mL or lower, four of the methods included an enrichment step.

**Table 3.** Positive detection of Ty21a in seeded samples measured via qPCR with a Ct ≤40

| Method<sup>a</sup> | CFU Ty21a/mL seeded |  |  |  |  |  |
| --- | --- | --- | --- | --- | --- | --- |
|  | 10,000 | 100 | 1 | 0.1 | 0.01 | 0.001 |
| FC1-D | 3/3 (100%) | 3/3 (100%) | - | - | 6/8 (75%) | 3/4 (75%) |
| FC2-D | 3/3 (100%) | 3/3 (100%) | - | - | 7/8 (88%) | 4/4 (100%) |
| DC-D-50mL | 3/3 (100%) | 3/3 (100%) | 6/6 (100%) | 6/6 (100%) | 3/6 (50%) | - |
| DC-SF-50mL | 3/3 (100%) | 3/3 (100%) | 0/6 (0%) | 4/6 (67%) | 1/6 (17%) | - |
| GE-UPE | 3/3 (100%) | 3/3 (100%) | 2/6 (33%) | 3/6 (50%) | 1/6 (17%) | - |
| MF1-D | 3/3 (100%) | 3/3 (100%) | 3/3 (100%) | 3/3 (100%) | 6/6 (100%) | 5/6 (83%) |
| MF1-OB | 3/3 (100%) | 3/3 (100%) | 3/3 (100%) | 2/3 (67%) | 6/6 (100%) | 1/6 (17%) |
| MF1-SC | 3/3 (100%) | 3/3 (100%) | 3/3 (100%) | 3/3 (100%) | 0/6 (0%) | - |
| MF2-SF | 3/3 (100%) | 3/3 (100%) | - | - | 6/6 (100%) | 2/6 (33%) |
| MS1-SF | 3/3 (100%) | 3/3 (100%) | 3/3 (100%) | - | 5/6 (83%) | 3/3 (100%) |
| MS2-UPE | 3/3 (100%) | 3/3 (100%) | - | - | 3/3 (100%) | 1/3 (33%) |

<sup>a</sup> naming convention shown in Table 1.
-, no data available.

In general, as the concentration of Ty21a seeded in the samples decreased, the Ct value increased linearly until the lowest Ty21a concentrations tested which had similar Ct values to the next lowest concentration (Figure S2).

Exceptions to this included MF1-OB, which had very low Ct values at a concentration 0.01 CFU/mL, and MS1-SF and MS-UPE, which had fairly consistent Ct values at all concentrations tested, likely due to the enrichment step utilized. With a seeded concentration of 10,000 and 100 CFU/mL, all samples yielded Ct values less than 40 with less than a 1-log variation in Ct values between replicates for a majority of sample types (82%, 9/11 and 73%, 8/11 for 10,000 and 100 CFU/mL, respectively).

### Ty2 detection

Five methods were further tested with Ty2: FC2-D, DC-D-1 L, MF1-D, MS1-SF, and MS2-UPE. When seeding 0.1 CFU Ty2/mL, four methods yielded high detection rates: FC2-D, DC-D-1L, MS1-SF, and MS2-UPE (>89%; Table 4). When the seeding concentration was decreased to 0.01 CFU/mL, DC-D-1L, MF1-D, MS1-SF, and MS2-UPE had high detection rates (>67%; Table 4). In general, seeding 0.1 CFU/mL resulted in a lower average Ct value and greater frequency of positivity when compared to seeding 0.01 CFU/mL (Figure S3).

**Table 4.** Positive detection of Ty2 in seeded samples measured via qPCR with a Ct ≤40

| Method<sup>a</sup> | CFU Ty2/mL seeded |  |
| --- | --- | --- |
|  | 0.1 | 0.01 |
| FC2-D | 11/12 (92%) | 3/6 (50%) |
| DC-D-1L | 8/9 (89%) | 7/9 (78%) |
| MF1-D | 6/18 (33%) | 6/6 (100%) |
| MS1-SF | 2/2 (100%) | 2/3 (67%) |
| MS2-UPE | 5/5 (100%) | 7/9 (78%) |

<sup>a</sup> naming convention shown in Table 1.

## DISCUSSION

### Ty21a and Ty2 detection

This study examined the detection and percent positivity of Ty21a by 11 ES concentration methods and Ty2 by 5 ES concentration methods. Methods were not optimized or evaluated for recovery efficiency, but rather chosen based on percent positivity and feasibility. As anticipated, when the concentration seeded into the wastewater decreased, the positive detection rate generally decreased. Exceptions to this were seen with decreased recovery of Ty21a at an intermediate concentration in FC1-D, FC2-D, and MS1-SF samples, which could be due to variability between experiments as the wastewater matrix was collected at different times for each experiment, and variability in the Ty21a strain as it is attenuated. Typically, methods that assayed larger volumes of the initial sample generally yielded a higher positive detection rate at low seeded concentrations than methods that assayed smaller volumes of seeded wastewater. A high percent recovery was measured at low Ty21a seeding concentrations for FC1-D and FC2-D methods, which assay the largest volume of the initial sample (85 and 123 mL, respectively; Table 1), and for MS1-SF samples. The Moore swab methods are inherently non-volumetric. The hold-up volume of the Moore swabs was approximately 60 mL (amount of liquid the Moore Swabs absorbed), and they were exposed to 5 L of recirculated sample in this study. Methods that assayed less than 1 mL of the original sample included DC-SF-50mL (minimal detection at 0.01 CFU Ty21a/mL), MF1-OB (minimal detection at 0.001 CFU Ty21a/mL), MF1-SC (no detection at 0.01 CFU Ty21a/mL), and GE-UPE (minimal detection at 1 and 0.1 CFU Ty21a/mL, and no detection at 0.01 CFU Ty21a/mL). This information was factored into the decision to exclude these methods from further experiments conducted with Ty2.

For all methods tested with Ty2, when the seeded concentration decreased, the positive detection rate decreased. Additionally, for methods with a calculable volume of the original sample assayed, high volumes processed yielded more positive detection, with DC-D-1L (9.7 mL) yielding the highest rate of positive detection, followed by FC2-D (123 mL), and MF1-D (2 mL). Finally, where the same assay was used, Ty21a was detected at lower seeded concentrations than Ty2. This could be due to differences in assay sensitivity to Ty21a and Ty2, variability in experiments or wastewater matrix used, differences in the organisms capture by these methods, or differences in the stability of these organisms in the wastewater matrix throughout processing.

### Method feasibility and use case scenarios

Method selection depends on field logistics, laboratory constraints, project design, and budgetary considerations; an appropriate ES method will operate within these confines while maintaining effective performance. Local field conditions and infrastructure impact appropriate surveillance sites (*e*.*g*., sewer, wastewater treatment plant pumping station, river, pit latrine, etc.) and therefore the appropriate matrix. These matrices will vary by available sample volume, total solids, and solid characteristics (*e*.*g*., sediments, debris), which in turn will inform appropriate sampling and concentration method selection. For example, matrices with high solids content may be a challenge for filter cartridge and membrane filtration methods due to filter clogging, while methods like GE-UPE, differential centrifugation, and Moore swabs are able to process samples with high solids contents. Filter cartridge methods rely on the adsorption of negatively charged bacteria onto positively charged filters with a pore size of 2-3 µm and thus these methods may be less applicable in turbid waters (*e*.*g*., high strength wastewater or pit latrine waste) as less volume may be able to be filtered which will affect recovery. Additionally, sampling sites with very low flows may not be applicable for methods that process large volumes such as filter cartridge, DC-D-1L, or membrane filtration.

Site access and field worker safety are critical considerations for sample collection and in-field processing. If sample shipment between the collection site and processing laboratory is required, then this may impact the sample volume (requiring smaller volume samples like with DC-D-50mL, DC-SF-5mL, or GE-UPE), the need to perform primary concentration at the field site (such as with the filter cartridge methods or the in-field version of Moore swabs), and/or the ability to maintain sample integrity over the period of transport. It is important to note that the Moore swabs methods tested in this evaluation were not representative of what happens in their intended use case as the experimental Moore swab was held in a recirculating system with multiple exposure to the same seeded wastewater rather than placed in a drainage with exposure only to new wastewater throughout the holding period.

The available laboratory equipment and supplies, physical space, and personnel time also impact the ability to conduct methods. For example, while the filter cartridge methods use a commercialized kit making procurement simple, they also require a large centrifuge and shaking table. Centrifuge capacity is also a challenge for DC-D-1L, particularly if the centrifuge does not contain a cooling mechanism or if multiple samples must be processed in one day. Access to a house vacuum or strong vacuum pump could also be a challenge preventing use of GE-UPE, membrane filtration methods, and MS2-UPE. Laboratorian safety is critical when choosing an appropriate ES method. Both Selenite Cysteine (used for MF1-OB and MF1-SC) and Selenite F (used for DC-SF-50mL, MF2-SF, and MS1-SF) broths must be prepared and used under a biosafety cabinet capable of chemical protection (*e*.*g*., Class II B2), due to their acute toxicity and teratogenicity. Selenite-based enrichment broths cannot be autoclaved and must be disposed as hazardous chemical waste due to their aquatic toxicity. Therefore, these chemical hazards are important considerations in settings where Selenite-based compounds cannot be contained or disposed of appropriately.

Finally, study design, time to results, and associated budgetary considerations necessarily impact method selection. In cases where results are needed rapidly, filter cartridge methods, DC-D-50mL, DC-D-1L, or MF1-D may be ideal as results can be obtained in less than 24 hours as no overnight incubation steps are required. This combination of field, laboratory, study, and performance considerations result in a complex decision tree where one outcome is not appropriate for all applications. Thus, it is necessary to have the flexibility to select an appropriate method that meets all or a majority of these needs. Here, different use case scenarios are outlined with potential appropriate methods (Table 5).

**Table 5.** Appropriate use cases for ES methods.

| Method name | TCV campaign location selection | TCV campaign monitoring | Low concentration detection | Disease surveillance |
| --- | --- | --- | --- | --- |
| FC1-D | X | X | X |  |
| FC2-D | X | X | X |  |
| DC-D-50mL | X |  | X | X |
| DC-SF-50mL | X* |  |  | X |
| DC-D-1L | X | X | X | X |
| GE-UPE | X |  |  | X |
| MF1-D | X | X | X | X |
| MF1-OB | X* |  |  | X |
| MF1-SC | X* |  |  | X |
| MF2-SF | X* | X* | X | X |
| MS1-SF | X* | X* | X | X |
| MS2-UPE | X* | X* | X | X |

X, indicates an appropriate method for the indicated use case.
X\*, indicates an appropriate method for the indicated use case with an increase in sample size or a most probable number approach.

#### TCV campaign location selection

ES data can be used by decision-makers for selecting high-burden locations to implement TCV campaigns using either qualitative or quantitative methods. If spread of Typhoid is anticipated to be high within a population, then effective volume assayed and sensitivity will be less critical to delineate true positive results, and results may be quantifiable. Whereas in lower prevalence areas, an understanding of the lower limit of detection is important to determine fit for purpose. Methods that involve an enrichment step could be utilized with a substantial increase in the sample number to improve understanding of the generated results. For this use case, all methods would be appropriate (Table 5).

#### TCV campaign monitoring

After a vaccine campaign is implemented, it is critical to continue ES as a quality control check on effective implementation by observing an anticipated decrease in the wild-type organism. A low concentration of wild-type *Salmonella* Typhi would be anticipated due to vaccine efficacy, therefore an ES method with high sensitivity and a high effective volume assayed would be appropriate. Additionally, a quantitative method (*e*.*g*., a discrete volume processed and no enrichment) could best inform on *Salmonella* Typhi presence before and after vaccine distribution. While a qualitative method could be used, this would require a substantial increase in sample number (via Moore swab) or splitting a sample and enriching at multiple input volumes, to see an effect via a most probable number approach. Appropriate methods may include filter cartridges, high volume differential centrifugation without enrichment, and membrane filtration without enrichment (Table 5).

#### Low concentration detection/Early outbreak detection

Prior to the outset of community spread, low or no concentrations of *Salmonella* Typhi would be anticipated in wastewater or wastewater-impacted surface waters. Consequently, an ES method aimed at early outbreak detection should have a high effective volume assayed and low limit of detection to look for a relative increase in concentration and percent positivity. Additionally, if a presence/absence result was adequate to alert health centers of an impending outbreak, then a qualitative ES method (*e*.*g*., non-discrete volume processed or enrichment) would be appropriate and may increase the likelihood of detection. Moore swabs allow a large volume to pass through the swab over time, and thus increase the potential for samples to collect the target organism. Enrichment increases the copy numbers of the target organism prior to assay via qPCR or culturing, subsequently increasing the potential to capture the organism in the volume assayed as well as informing on the viability of enriched bacteria. Potential methods include filter cartridges, differential centrifugation, membrane filtration, and Moore swabs (Table 5). This use case is aspirational as it would require routine surveillance to be successful.

#### Disease surveillance

Routine surveillance to monitor geographic disease spread within the context of an active outbreak could be conducted using either qualitative or quantitative methods. In poorly resourced health systems, ES might also act as a trigger to scale up bacteremia surveillance, to confirm the etiology of the outbreak and establish the antimicrobial susceptibility of the strain causing the outbreak. An active outbreak would indicate higher anticipated *Salmonella* Typhi concentrations, and so method sensitivity may be less critical in these instances. Further, routine or intense targeted surveillance can be costly and so incorporating a less expensive method that still yields results may be a sustainable solution. Appropriate methods may include differential centrifugation, grab enrichment, membrane filtration, and Moore swabs (Table 5).

## Limitations

Limitations of this study included variability between and within experiments, the number of replicates for different methods and seeding levels evaluated, the assay chosen for detection, and the methods evaluated. Variability between experiments could result from the wastewater matrix used. While the background matrix was collected from the same local wastewater treatment plant for every experiment, the experiments were conducted over a 1-year timeframe due to laboratory capacity to process samples. Additionally, wastewater could not be collected once and stored for all experiments as wastewater characteristics change with storage time and limited storage space was available. Thus, there may have been variability in the wastewater matrix composition due to precipitation events prior to or during certain sample collections, as the treatment plant is a combined sewer overflow system.

Additionally, while glycerol stocks of the same initial culture of Ty21a or Ty2 were used to prepare the organisms for seeding, and growth curves were determined, there was minor variation in the length of the overnight cultures, potentially introducing variability in the anticipated amount of Ty21a or Ty2 seeded. Overnight cultures were quantified via spread plating with differences from the anticipated amount ranging from 8 to 156% for Ty21a and 11 to 498% for Ty2. There could also be variability within an experiment as it is difficult to fully homogenize large sample volumes. The effect of this could be amplified at low seeding levels. While the extraction method used was consistent across all sample types, the extraction efficiency of the different sample types could vary. Finally, when Selenite F and Selenite Cysteine broths were used, the length of the overnight culture in the selenite broths varied from 12-18 hours and 20-21 hours. Longer enrichment in selenite-based media can overtax the media leading to breakthrough of organisms other than the target, *Salmonella* Typhi.

Variability in the number of replicates per method and per experiment makes comparisons across methods and seeding levels challenging, and only a limited number of replicates were generated for the methods evaluated. The differences in the number of replicates for the different methods were due to the complexity and feasibility of processing multiple replicates using a certain method, as some methods required less wastewater or personnel time than others. For example, the large variability in Ct values seen at 0.01 CFU Ty2/mL compared to 0.1 CFU Ty2/mL with DC-D-1L could be due to the differences in number of experiments (three were conducted at 0.01 CFU/mL and one conducted at 0.1 CFU/mL), number of replicates, and challenges with homogenizing a low concentration of Ty2 throughout a sample. Additionally, it is expected to see large variation between replicates at low concentrations that approach the assay limit of detection.

This study utilized the Nga *et al*. primer and probe set for detection of *Salmonella* Typhi. This assay was originally designed for clinical samples and has recently been applied for detection in environmental samples. However, wastewater contains a variety of organisms and both known and unknown DNA in the samples, which could interfere with this assay and result in cross reactivity [44]. There may be variability between concentration methods in the ability to detect *Salmonella* Typhi using this assay, as some methods concentrate larger volumes resulting in greater concentration of potential qPCR inhibitors. To minimize these effects on reported results, undiluted and 10-fold sample dilutions were assayed to screen for evidence of inhibitors. Future studies should examine improved qPCR assays for environmental surveillance and archived samples from this study could be retested with the optimized method.

Finally, this study evaluated multiple *Salmonella* Typhi ES methods, though some other ES methods were not able to be included. For example, additional ES methods such as dead-end ultrafiltration and hollow fiber ultrafiltration were not able to be included in this study due to time and resource constraints. Future studies should expand on the types of methods evaluated.

## CONCLUSIONS

This study evaluated eight methods with twelve formats for their applicability to conduct ES using *Salmonella* Typhi Ty21a and Ty2. Results suggest that all methods tested can be successful at concentrating *Salmonella* Typhi for subsequent detection by qPCR, although each method has its own strengths and weaknesses including the *Salmonella* Typhi concentrations they are applicable for use with. These factors should be considered when identifying a method for *Salmonella* Typhi ES and will greatly depend on the use case planned. Future studies could benefit from examining additional ES methods not used here and conducting side-by-side evaluations with field samples.

## Data Availability

All data produced in the present study are available upon reasonable request to the authors

## Notes

### Competing Interest Statement

The authors have declared no competing interest.

### Funding Statement

Funding was provided by the Bill and Melinda Gates Foundation, Grant Numbers INV-000508 and INV-026705. NAZ was further supported in part by the UW NIEHS sponsored Biostatistics, Epidemiologic and Bioinformatic Training in Environmental Health (BEBTEH) Training Grant, Grant #: NIEHS T32ES015459.

