## Supplementary Information for "Evaluation of sampling and concentration methods for *Salmonella enterica* serovar Typhi detection from wastewater"

**Short title:** Evaluation of *Salmonella* Typhi environmental surveillance methods

**Author names**

Nicolette A. Zhou, Angelo Q.W. Ong, Christine S. Fagnant-Sperati, Joanna Ciol Harrison, Alexandra L. Kossik, Nicola K. Beck, Jeffry H. Shirai, Elisabeth Burnor, Rachael Swanstrom, Bethel Demeke, Suhani Patel, Typhoid Environmental Surveillance Working Group, John Scott Meschke*

**Corresponding Author**

*corresponding author:; phone: +1-206-221-5470; postal: Department of Environmental and Occupational Health Sciences, University of Washington; 4225 Roosevelt Way NE, Suite 100, Seattle, WA 98105, USA

**Methods evaluated**

*Two-inch filter cartridge method*

Two variations of the two-inch filter cartridge method were tested, mentioned hereafter as FC1-D and FC2-D. For both versions, seeded wastewater was filtered through ViroCap filters (Scientific Methods Inc., Granger, Indiana, USA) as described previously [19,20] until 40 minutes or 3 L had passed, whichever took longer (Table 1). The volume filtered was recorded. Once filtered, the ViroCap filters were eluted via a double elution as previously described with an input volume of 150 mL, for a final eluent volume of 300 mL [39]. Secondary concentration was performed on the eluates via skimmed milk flocculation [39]. Skimmed milk was added to the eluates and the pH was lowered to 3.5-4.0 using HCl and NaOH as needed. Samples were shaken (200 rpm, 2 hours, ambient temperature), centrifuged (3500 ×*g*, 30 minutes, 4°C), and the supernatant was discarded. For FC1-D samples, the secondary concentrate pellet from 150-mL of eluate was resuspended into 2-mL 1×PBS. One (1)-mL aliquots of this resuspension were then centrifuged (10,000 ×*g*, 10 minutes, ambient temperature), the supernatant discarded, and the pellet stored for DNA extraction (-20°C). For FC2-D samples, the secondary concentrate pellet from 150 mL of eluate was stored at -20°C prior to DNA extraction.

*Differential centrifugation*

Three versions of differential centrifugation methods were used: DC-D-50mL, DC-SF-50mL, and DC-D-1L. Differential centrifugation samples (50mL or 1L volumes) were divided into 50 mL conical tubes and centrifuged (1 minute, 1000 ×*g*, 4°C). The supernatant from each conical was transferred to a new conical tube and centrifuged again (15 minutes, 4000 ×*g*, 4°C). The supernatant was then discarded and all pellets combined into 1.5 mL (for DC-D-50mL samples) or 4 mL (for DC-D-1L samples) 1×PBS. Lastly, the resuspended sample was vortexed, aliquoted into 1 mL volumes, centrifuged (10,000 ×*g*, 10 minutes, ambient temperature), the supernatant discarded, and the pellet stored at -20°C until DNA extraction. DC-SF-50mL samples were prepared by transferring 0.5 mL of the resuspended DC-D-50mL sample into a 15 mL conical tube, adding 6 mL Selenite F broth (HiMedia, Chester, PA, USA), and incubating with the lid loosely capped (20-21 hours, 37°C). After incubation, the enrichment was distributed into 1 mL volumes, centrifuged (10,000 ×*g*, 10 minutes, ambient temperature), supernatant discarded, and stored at -20°C prior to DNA extraction.

*Membrane filtration methods*

Multiple variations of vacuum membrane filtration were tested: MF1-D, MF1-OB, MF1-SC, and MF2-SF (Fig. 1). After distribution of seeded wastewater samples (1L each), MF1-D, MF1-OB, and MF1-SC samples were gently swirled to resuspend solids and added to a paper, cone-shaped coffee filter placed on top of a filtration cup with an MCE filter. The coffee filter acted as a pre-screen for larger solids. Due to filter clogging, the coffee and MCE filters were replaced each hour, until five filters were obtained per sample. The remaining unfiltered volume was measured, and total volume filtered recorded (Table 1). The five filter discs were then transferred to a Whirl-Pak® bag and 10 mL Ringer's lactate solution (Alfa Aesar, Haverhill, MA, USA) added. The filter discs were massaged in the bags until the filters appeared clean or broken apart. The eluate was then processed in three ways prior to DNA extraction: unenriched eluates (MF1-D); Ox bile (Sigma Aldrich) to Selenite Cystine (Becton, Dickinson and Company) enrichments (MF1-OB); and Selenite Cystine to Ox bile enrichments (MF1-SC).

For MF1-D samples, two 1 mL volumes from the eluate were collected, centrifuged (10,000 ×*g*, 10 minutes, ambient temperature), the supernatant discarded, and the pellet stored at -20°C prior to DNA extraction. For MF1-OB samples, 1 mL of the eluate was added to 9 mL of Ox Bile broth and incubated (18-24 hours, 37°C). After incubation, 5 mL of the enrichment was added to 5 mL of 2×Selenite Cystine broth and incubated (12-18 hours, 41°C). The enrichment was then distributed into 1 mL volumes, centrifuged (10,000 ×*g*, 10 minutes, ambient temperature), supernatant discarded, and stored at -20°C prior to DNA extraction. For MF1-SC samples, 5 mL of the eluate was added to 5 mL of 2×Selenite Cystine broth and incubated (12-18 hours, 41°C). After incubation, 1 mL of Selenite Cystine enriched eluate was added to 9 mL of Ox Bile broth and incubated (18-24 hours, 37°C). The enrichment was then distributed into 1 mL volumes, centrifuged (10,000 ×*g*, 10 minutes, ambient temperature), supernatant discarded, and stored at -20°C prior to DNA extraction.

After distribution of seeded wastewater samples, MF2 samples were swirled to mix and added directly on an MCE filter (no coffee filter pre-screen was used). Each sample was filtered for one hour with the entire process repeated until six filters were obtained per sample. The membrane filters were then transferred to a conical, 10 mL Selenite F broth was added, and the sample incubated with the lid loosely capped (16-17 hours, 37°C). After incubation, the enrichment was distributed into 1 mL volumes, centrifuged (10,000 ×*g*, 10 minutes, ambient temperature), supernatant discarded, and stored at -20°C prior to DNA extraction.

*Moore swab methods*

Two Moore swab methods were tested, with one enriched using Selenite F (SF) broth (hereafter, MS1-SF) and another using UPE broth (hereafter, MS2-UPE). The Moore swabs were made from 6 feet (length) by 1.5 feet (width) of sterile hospital gauze by accordion-folding the gauze width-wise with a fold width of 1 inch and tying a nylon string in the middle [28]. To simulate the usage of the Moore swabs in the environment, a recirculating pumping system was devised, in which a Moore swab was placed in 5 L of seeded wastewater in a 10-L carboy with a ported lid. Sterile tubing fed from the carboy through a peristaltic pump, and back into the carboy, for continuous circulation with laminar flow to simulate water flow through a sewage conveyance line or a river system. The wastewater was recirculated for 72 (MS1-SF) or 24 hours (MS2-UPE) prior to swab removal from the carboy.

The swab was placed into a sterile bottle and enriched in Selenite F broth (200 mL) or UPE broth (450 mL) overnight (37°C). MS1-SF samples were collected after incubation by aliquoting 1 mL volumes of the enrichment, centrifuging (10,000 ×*g*, 10 minutes, ambient temperature), discarding the supernatant, and storing the pellet at -20°C prior to DNA extraction. After incubation, MS2-UPE samples were collected by performing membrane filtration on a 20 mL volume of the enrichment. The membrane filter was sliced into 6-10 pieces using sterilized scissors and placed in a 2 mL screw top tube and stored at -20°C prior to DNA extraction.

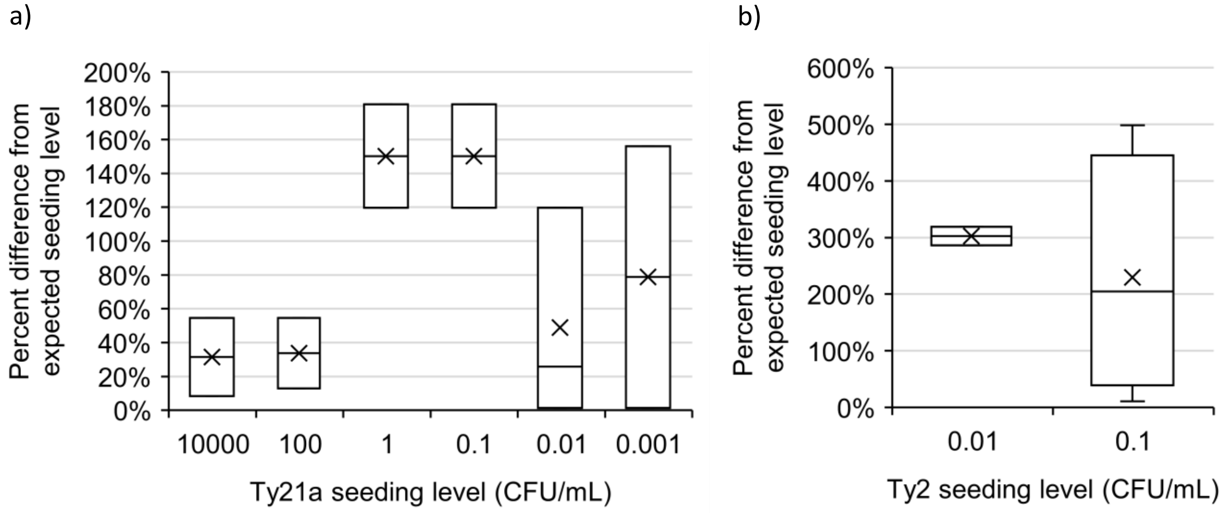

**Figure S1.** Percent difference between the a) Ty21a and b) Ty2 concentration seeded from the expected seeding level.

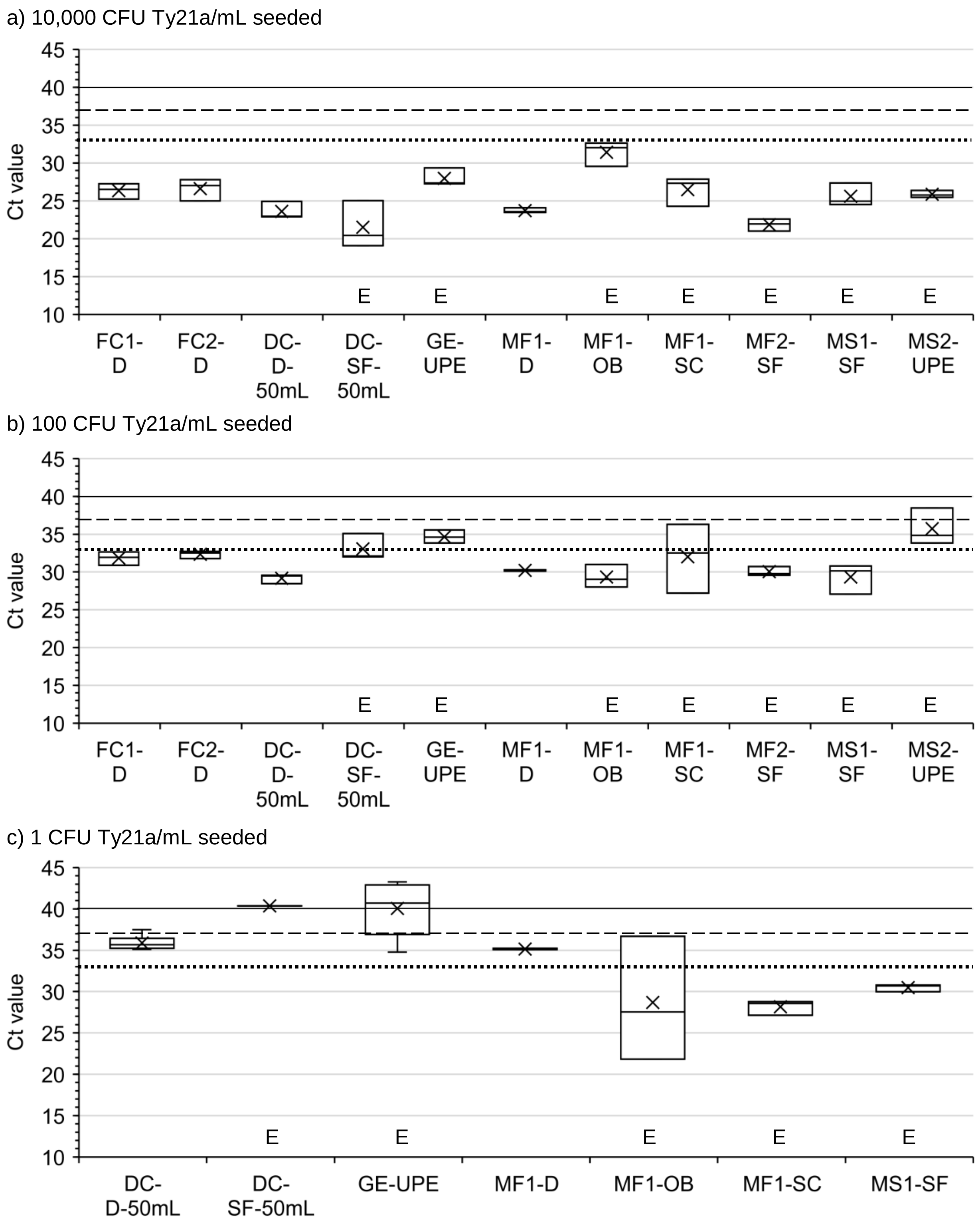
a) 10,000 CFU Ty21a/mL seeded
Ct value
45
40
35
30
25
20
15
10
E E E E E E E
FC1-D
FC2-D
DC-D-50mL
DC-SF-50mL
GE-UPE
MF1-D
MF1-OB
MF1-SC
MF2-SF
MS1-SF
MS2-UPE
b) 100 CFU Ty21a/mL seeded
Ct value
45
40
35
30
25
20
15
10
E E E E E E E
FC1-D
FC2-D
DC-D-50mL
DC-SF-50mL
GE-UPE
MF1-D
MF1-OB
MF1-SC
MF2-SF
MS1-SF
MS2-UPE
c) 1 CFU Ty21a/mL seeded
Ct value
45
40
35
30
25
20
15
10
E E E E E
DC-D-50mL
DC-SF-50mL
GE-UPE
MF1-D
MF1-OB
MF1-SC
MS1-SF

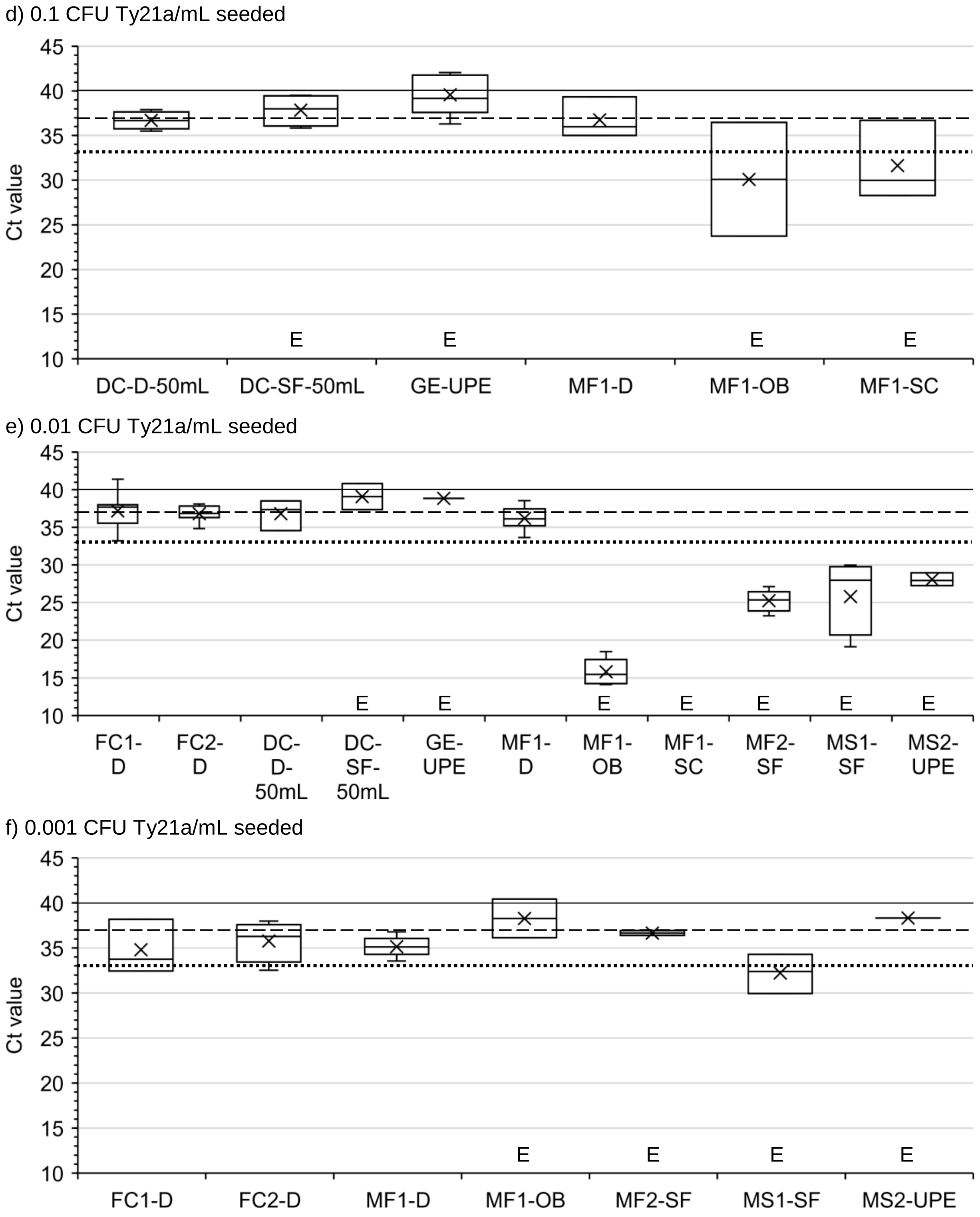

**Figure S2.** Box and whisker plots of Ty21a detection in samples seeded with a) 10,000, b) 100, c) 1, d) 0.1, e) 0.01, and f) 0.001 CFU Ty21a/mL and processed by various methods. Upper, middle, and lower box lines show the first, second, and third quartiles, respectively; whiskers show the minimum and maximum data points; markers 'x' show the mean; 'E' indicates there is an enrichment step present in the method. Method naming convention shown in Table 1. Samples below Ct 40 were considered positive (solid horizontal line shown). Limit of detection shown by dashed horizontal line. Limit of quantification shown by dotted horizontal line.

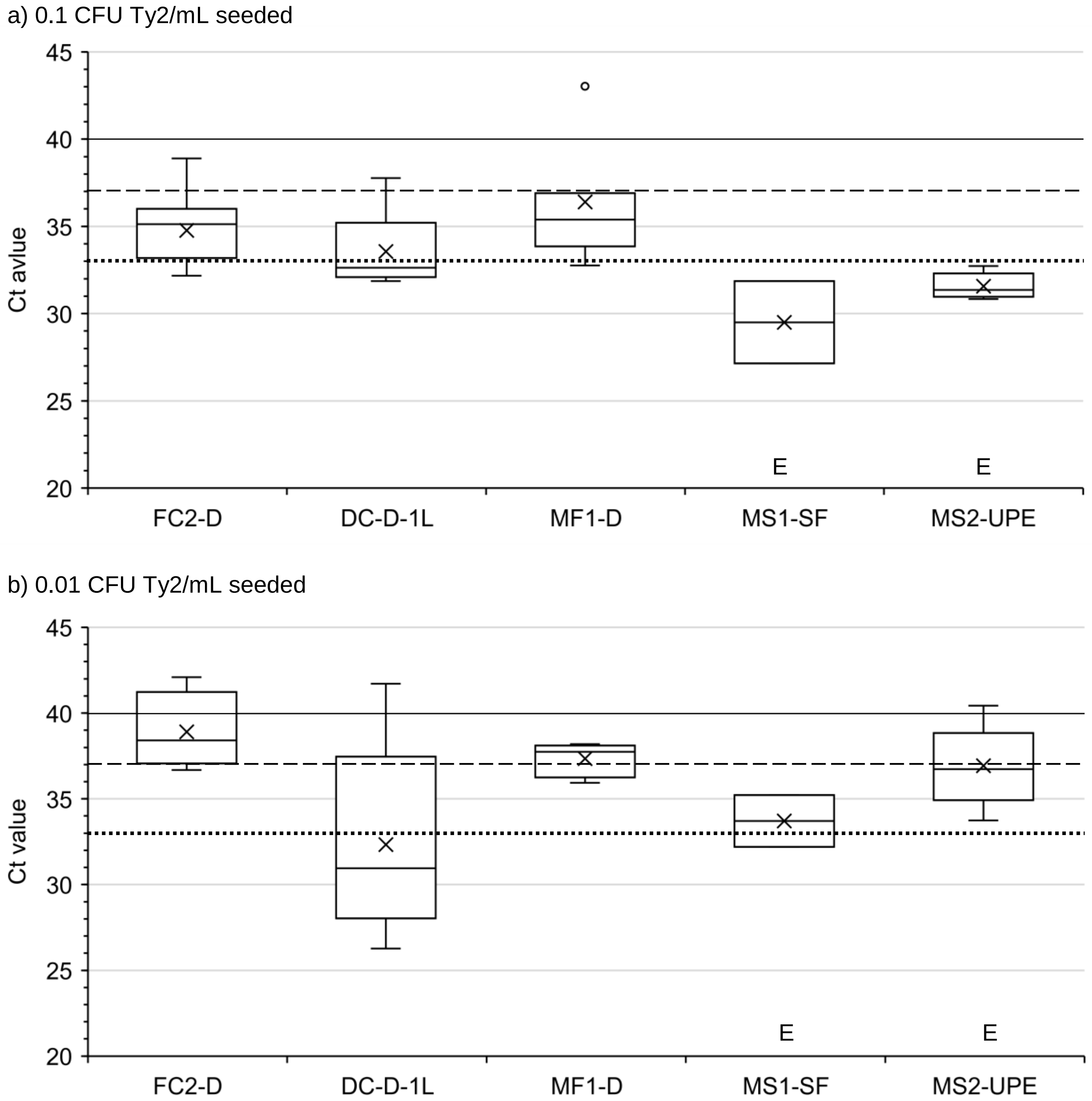

**Figure S3.** Box and whisker plots of Ty2 detection in samples seeded with a) 0.1 and b) 0.01 CFU Ty2/mL and processed by various methods. Upper, middle, and lower box lines show the first, second, and third quartiles, respectively; whiskers show the minimum and maximum data points; markers 'x' show the mean; 'E' indicates there is an enrichment step present in the method. Method naming convention shown in Table 1. Samples below Ct 40 were considered positive (solid horizontal line shown). Limit of detection shown by dashed horizontal line. Limit of quantification shown by dotted horizontal line.
